# Intraoperative Very High-Frequency Activity for Localization of the Epileptogenic Zone: An International Multicenter Study

**DOI:** 10.64898/2026.10.14.26365543

**Authors:** Hiroshi Uda, Naoto Kuroda, Naotaka Usui, Hiroshi Ogawa, Taro Okumura, Mitsuru Hashiguchi, Norihiko Kawaguchi, Keisuke Hatano, Ece Erder, Keitaro Fujioka, Dominik Choromanski, Naznin Mahmood, Michelle Figueroa, Norimitsu Kuwabara, Aimee F. Luat, Michael Cools, Eishi Asano

## Abstract

**Background and Objectives:** Interictal spikes on intraoperative electrocorticography have been used to localize the epileptogenic zone but have limited specificity. We hypothesized that identifying spatially distributed, spike-associated very high-frequency activity (vHFA; 500–2,000 Hz) would improve the ability of spikes to localize the epileptogenic zone. We aimed to characterize spike-associated vHFA and to determine whether a seizure outcome prediction model incorporating vHFA could be externally validated across independent centers.

**Methods:** This retrospective two-center cohort study included patients with drug-resistant focal epilepsy who underwent electrocorticography and subsequent resection between January 2016 and May 2025. Spikes occurring across contacts within 50 ms were classified as co-occurring; all others were classified as isolated. We determined whether vHFA was associated with co-occurring spikes. We then assessed whether contacts enriched in vHFA-positive co-occurring spikes were more likely to be located within the resected region among seizure-free patients. Finally, we determined whether the rate of such spikes remaining outside the resection predicted seizure outcomes across the two centers.

**Results:** Among 56 patients (mean age: 21.2 years; 26 females), we identified 132,359 spikes, of which 1,925 were vHFA-positive. The vHFA peak preceded the associated spike peak by 8.0 ms (95%CI: 5.0–16.0). vHFA-positive spikes were more likely to be classified as co-occurring, even after adjustment for anatomy and inter-electrode distance (OR: 3.43; 95%CI: 2.82–4.17). Among the 35 seizure-free patients, contacts with higher rates of vHFA-positive co-occurring spikes were more likely to be resected (OR: 11.92 per occurrence/min; 95%CI: 5.62–25.32). This association was stronger than that observed for all spikes (OR: 1.09 per occurrence/min; 95%CI: 1.06–1.11). Twenty patients experienced postoperative seizure recurrence; a higher proportion of vHFA-positive co-occurring spikes in preserved areas was associated with recurrence (OR: 3.22 per 0.01 increase in proportion; 95%CI: 1.14–9.12). In bidirectional cross-center validation, models incorporating vHFA-positive co-occurring spikes yielded areas under the receiver operating characteristic curves of 0.80 and 0.69, with better performance when the model was developed using the larger sample.

**Discussion:** Spike-associated vHFA may identify a biologically meaningful subset of interictal spikes involved in propagation across the epileptogenic zone. vHFA-positive co-occurring spikes may represent a generalizable intraoperative biomarker of the epileptogenic zone.

## Introduction

Accurate localization of the epileptogenic zone is fundamental to achieving seizure freedom after resective surgery for drug-resistant focal epilepsy. Intraoperative electrocorticography (ECoG) has traditionally relied on interictal spikes as electrophysiologic markers, partly because spikes can be captured during relatively brief recordings.^1–3^ Automated spike detection has been employed to improve interrater reproducibility; however, the localizing value of interictal spikes remains limited by their suboptimal specificity.^4^ Consequently, investigators have sought characteristics that distinguish spikes arising from the epileptogenic zone.

Investigators have proposed that the *spectral* characteristics of spike discharges, particularly spike-associated high-frequency activity (HFA) at ≥80 Hz, may improve their specificity for the epileptogenic zone. Studies of extraoperative intracranial EEG (iEEG) have reported that spikes coupled with HFA at 80–250 Hz^5^ or 250–500 Hz^6^ may be more specific markers than spikes alone, as incomplete resection of brain tissue exhibiting such events was associated with poorer postoperative seizure outcomes. Very high-frequency activity (vHFA) at 500–2,000 Hz can also be recorded using intracranial macroelectrodes and has been proposed as a marker of the epileptogenic zone.^7,8^ In one study, ictal vHFA was visually identified in 7 of 13 patients undergoing extraoperative iEEG, and its presence was associated with favorable postoperative seizure outcomes.^7^ In another study, interictal vHFA was detected in 23 of 40 patients; among the 10 patients included in the resection–outcome analysis, resection of a greater proportion of vHFA-positive contacts was associated with better postoperative seizure outcomes.^8^ However, the prognostic value of vHFA in intraoperative ECoG recordings remains unclear.

The *spatiotemporal* characteristics of spike discharges may likewise enhance their specificity for the epileptogenic zone. Spikes that co-occur across multiple recording sites within tens of milliseconds, particularly those occurring earlier in the propagation sequence, have been proposed to better localize the epileptogenic zone.^1,9–11^ A study of 41 patients undergoing extraoperative iEEG reported that failure to resect the onset regions of propagating spikes coupled with HFA_80– 250 Hz_ was associated with poor postoperative seizure outcomes.^12^ Whether spatiotemporal co-occurrence and spike-associated vHFA jointly enhance the localizing value of spikes remains unclear.

Here, we investigated the localizing and prognostic value of spike-associated vHFA detected using macroelectrodes. We employed time-frequency analysis of intraoperative ECoG to objectively define and detect vHFA-positive spikes and characterize their spatiotemporal co-occurrence patterns. We then evaluated the utility of vHFA-positive co-occurring spikes for predicting postoperative seizure outcomes and assessed whether the resulting prediction models could be externally validated between centers. Internal validation, including leave-one-out cross-validation, has often been employed to assess the utility of electrophysiological biomarkers; however, such approaches may yield overly optimistic estimates of predictive performance and cannot establish generalizability across centers.^13–15^ External validation is essential to establish the generalizability of proposed electrophysiological biomarkers,^16–19^ given intercenter differences in patient demographics, epilepsy etiology, electrode placement, and anesthetic conditions. Biomarkers that remain predictive despite such differences have *greater* clinical utility. We hypothesized that vHFA-positive spikes would preferentially co-occur across recording sites and that preservation of vHFA-positive co-occurring spikes would predict postoperative seizure recurrence in both cohorts.

## Methods

### Patients

We conducted a retrospective, two-center observational cohort study. Patients were eligible if they (1) underwent curative focal resection for drug-resistant focal epilepsy at Children’s Hospital of Michigan in Detroit, USA (Detroit), or the National Epilepsy Center, NHO Shizuoka Institute of Epilepsy and Neurological Disorders in Shizuoka, Japan (Shizuoka), between January 2016 and May 2025; (2) had intraoperative ECoG recorded at 10,000 Hz under sevoflurane anesthesia; (3) had intraoperative photographs or videos available to determine electrode locations and the extent of resection (Figure 1A)^17^; and (4) had at least 1 year of postoperative follow-up and an outcome classified according to the International League Against Epilepsy (ILAE) outcome scale.^20^ Patients were excluded if they had previously undergone resective epilepsy surgery.

**Figure 1.**
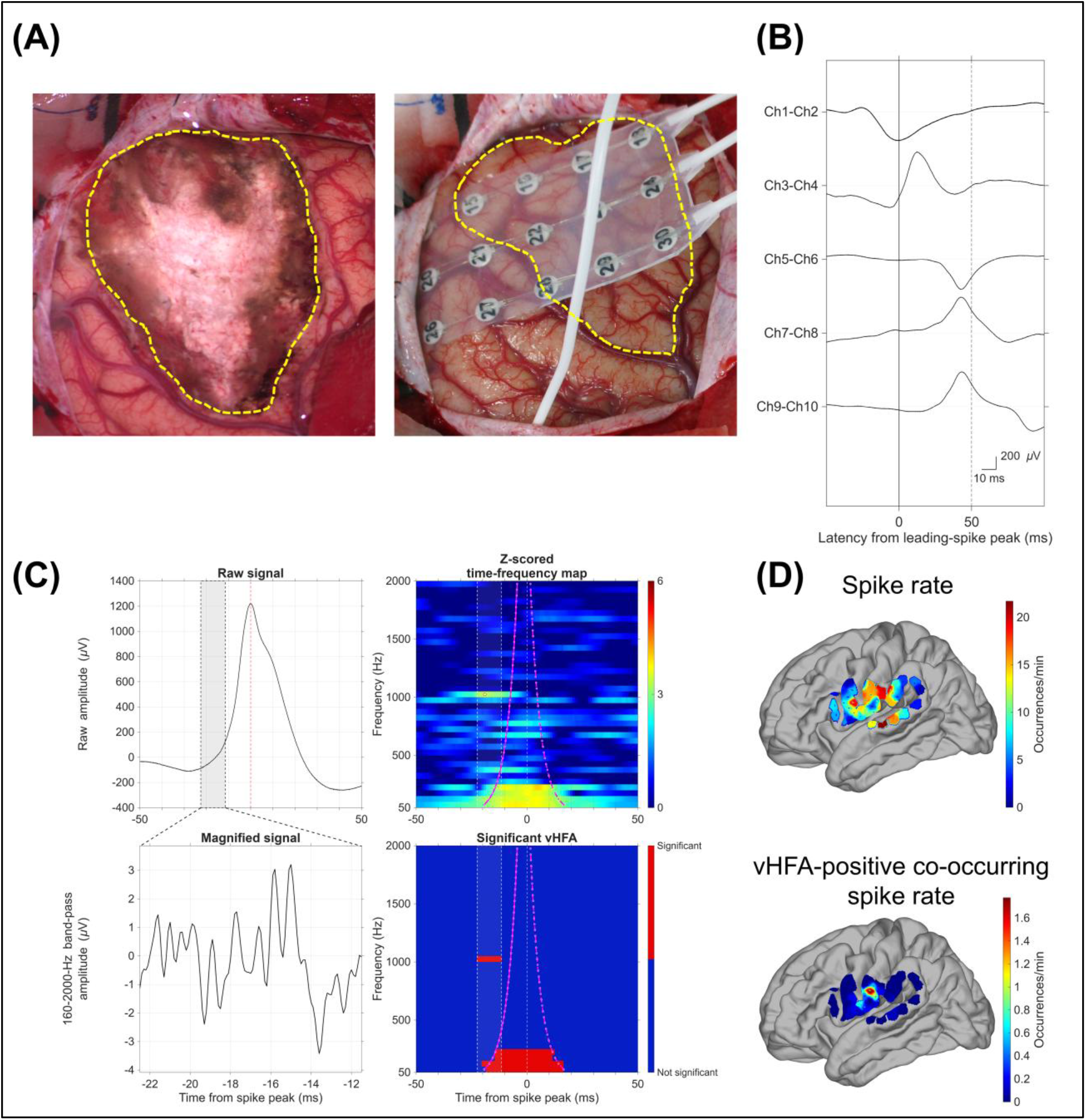
Summary of the analysis workflow. **(A)** Intraoperative photographs show the intracranial electrodes used for electrocorticography (ECoG) and the subsequent resection margins (yellow dashed lines). **(B)** Co-occurring spike events within the 50-ms window. Channel 1–2 shows the leading spike (downward deflection at 0 ms). Subsequent spikes within this window were classified as co-occurring lagging spikes. **(C)** Very high-frequency activity (vHFA) identified by time-frequency analysis. Magnified ECoG signal, band-pass filtered at 160–2,000 Hz, visualizes high-frequency activity during the rising phase of the spike. The vHFA begins before the expected temporal boundary of broadband spectral activity generated by sharp spike transients and filtering (magenta curved lines). The vertical line at 0 ms, and the white circle marks the vHFA peak latency. **(D) P**er-minute occurrence rates of all detected spikes (upper) and vHFA-positive co-occurring spikes (lower) are mapped onto the cortical surface of a patient who became seizure-free after resection of the left inferior postcentral gyrus.

### Intraoperative ECoG recording

We recorded intraoperative ECoG before surgical resection at a sampling frequency of 10,000 Hz with a hardware bandpass of 0.016–3,000 Hz (Neurofax; Nihon Kohden, Tokyo, Japan). General anesthesia was maintained with sevoflurane at concentrations ranging from 1 to 4 vol%, and the end-tidal carbon dioxide partial pressure was maintained below 40 mm Hg. In Detroit, intraoperative ECoG was recorded at multiple sevoflurane concentrations using stepwise adjustments to reduce the risk of failing to capture interictal spike, with continuous communication between the clinical neurophysiologist and anesthesiologist.^21,22^ For patients who underwent one-stage surgery (i.e., resection immediately following ECoG), we used platinum subdural electrodes (10-mm center-to-center spacing). For patients who underwent two-stage surgery with extraoperative iEEG following ECoG, we used subdural electrodes alone, a combination of subdural and depth electrodes (5-mm center-to-center spacing), or stereoencephalography (sEEG) depth electrodes (3.5-mm center-to-center spacing). Recorded ECoG data were imported using FieldTrip (https://www.fieldtriptoolbox.org) for subsequent electrophysiological analysis.

### Electrode localization

For patients undergoing one-stage surgery, we co-registered intracranial electrode locations with preoperative MRI using intraoperative photographs and videos obtained through the surgical microscope.^21,23,24^ For patients undergoing two-stage surgery, we co-registered electrode locations with preoperative MRI using post-implantation CT images,^25^ with localization accuracy confirmed using intraoperative photographs. Using FreeSurfer scripts (https://surfer.nmr.mgh.harvard.edu), we mapped all electrode sites onto the standard brain template^26,27^ and classified a bipolar contact pair as being in the medial temporal region if at least one of its constituent contacts was localized to the entorhinal, parahippocampal, or fusiform region (**eFigure 1**).

### Postoperative seizure outcome and operational definition of the epileptogenic zone

Postoperative seizure outcomes were classified according to the ILAE outcome scale at least 1 year after surgery.^20^ ILAE class 1 was considered seizure freedom; all other classes were considered postoperative seizure recurrence. For localization analyses restricted to patients with an ILAE class 1 outcome, the resected region served as the operational reference for the epileptogenic zone.

### Spike detection and subtypes

Electrodes exhibiting persistent artifacts on visual review were excluded from the analysis. Interictal spikes were detected on a bipolar montage as an amplitude augmentation in 10–60 Hz activity using an open-source automated detector with its default settings.^28^ The spike peak was defined as the largest voltage deflection in the ECoG signal sampled at 10,000 Hz. When multiple spikes occurred in the same contact within 2 seconds, only the first was retained to minimize time-frequency effects from subsequent spikes.

Spike events were classified as co-occurring when another spike occurred at a different contact within ±50 ms (**Figure 1B**) and otherwise as isolated. Among co-occurring spikes, the first event was classified as the leading spike and subsequent events as lagging spikes.^10,29^

### Time-frequency analysis and detection of very high-frequency activity (vHFA)

We identified and characterized vHFA_500–2,000 Hz_ within ±200 ms of each spike (**Figure 1C**) using the Signal Processing Toolbox in MATLAB R2025a (MathWorks, Natick, MA, USA). For each 50-Hz frequency bin, a fourth-order Butterworth bandpass filter was applied in a zero-phase manner using the ‘butter’ and ‘filtfilt’ functions. The analytic signal was obtained using the ‘hilbert’ function, and instantaneous power was calculated as its squared magnitude. Power ratios were quantified in 1-ms bins relative to mean power during the −1,800- to −1,000-ms pre-spike baseline.^30^ To identify *significant* power augmentation, log power ratios at each time-frequency point were z-standardized using an empirical surrogate distribution generated from 200 randomly timed epochs from the same bipolar channel. Candidate activity was defined as a z score ≥3 sustained for ≥5 consecutive bins within the same frequency bin.

Furthermore, to distinguish vHFA from broadband spectral activity generated by sharp spike transients and filtering, we estimated the expected temporal boundary of spike-related spectral activity from lower frequencies (**Figure 1C**). Specifically, the onset and offset times of significant activity at 50–250 Hz were fitted separately as linear functions of log-transformed frequency and extrapolated to higher frequencies. Candidate activity at 500–2,000 Hz was classified as *significant* vHFA only when it extended beyond the extrapolated onset or offset boundary (**Figure 1C**). Only significant vHFA was retained for subsequent analyses, and spikes with significant vHFA were designated vHFA-positive. Based on their spatiotemporal features, vHFA-positive spikes were classified as either co-occurring or isolated, with co-occurring spikes further classified as leading or lagging.

### Statistical analysis

Statistical analyses were performed using the Statistics and Machine Learning Toolbox in MATLAB R2025a. Binary outcomes were evaluated using logistic regression or mixed-effects logistic regression, with random intercepts specified for each analysis. Results are reported as odds ratios (ORs) with 95% confidence intervals (95%CIs). Discrimination was quantified using the area under the receiver operating characteristic curve (AUROC) and the area under the precision-recall curve (AUPRC), calculated using the MATLAB ‘perfcurve’ function; AUROCs were compared using DeLong tests. Bootstrap analyses used 10,000 resamples that preserved the sampling structure specified for each analysis. All tests were two-sided, with statistical significance defined as a Bonferroni-adjusted P value below 0.05 within each prespecified test family.

### Temporal features of vHFA

Using patient-level median peak latencies, we tested whether vHFA peak latency relative to the associated spike peak differed from zero using an exact sign test.

### Factors associated with vHFA

We aimed to identify factors associated with the presence of vHFA_500–2,000 Hz_ using spike event-level mixed-effects logistic regression with a patient random intercept. Separate univariable models evaluated whether co-occurring spikes were more frequently associated with the presence of vHFA than isolated spikes and whether leading spikes were more frequently associated with vHFA than lagging spikes. Separate univariable models likewise evaluated associations of vHFA with √age, sex, number of antiseizure medications, presence of daily seizures, sevoflurane concentration, preoperative MRI lesion status, medial temporal sampling, and study center. Because 10 separate tests were performed, unadjusted P values below 0.005 were considered statistically significant after Bonferroni correction.

### Association between vHFA and spatial co-occurrence of spikes

We aimed to determine whether the presence of vHFA was associated with higher odds of a given spike co-occurring with spikes on other contacts, after adjusting for medial temporal sampling and interelectrode distance. We fitted a multivariable, spike event-level mixed-effects logistic regression model with a patient random intercept, with spike co-occurrence as the outcome, vHFA positivity as the predictor, and medial temporal sampling and median interelectrode distance as covariates.

### Association between vHFA and the epileptogenic zone

We aimed to determine whether, among patients who achieved ILAE class 1 outcomes, contacts with higher rates of vHFA-positive co-occurring spikes were more likely to be located within the resected region (i.e., the presumed epileptogenic zone). We fitted a contact-level mixed-effects logistic regression model with a patient random intercept, with resection status as the outcome and the occurrence rate (events/min) of vHFA-positive co-occurring spikes as the predictor. As control analyses, we repeated the analysis using the occurrence rates of vHFA-positive leading spikes, vHFA-positive spikes, co-occurring spikes, and all detected spikes as predictors. Furthermore, to assess the utility of HFA_250-500 Hz_, we repeated the analysis using the occurrence rate of HFA_250-500 Hz_-positive co-occurring spikes as the predictor. For these six tests, P values below 0.0083 were considered statistically significant after Bonferroni correction.

### Association between vHFA and postoperative seizure recurrence

We aimed to determine whether a higher burden of vHFA-positive co-occurring spikes in preserved areas was associated with a higher risk of postoperative seizure recurrence using logistic regression. The predictor was the proportion of vHFA-positive co-occurring spikes outside the resected region relative to all detected spikes. The outcome was postoperative seizure recurrence, defined as ILAE class 2 or worse. As control analyses, we repeated the analysis using the corresponding proportions of each predictor described above.

### Prediction of postoperative seizure recurrence

We assessed the generalizability of vHFA-positive co-occurring spikes as a biomarker for predicting postoperative seizure recurrence using bidirectional cross-center external validation. We first developed a logistic regression model using data exclusively from one center and evaluated its predictive performance in the independent cohort from the other center. The predictor was the proportion of vHFA-positive co-occurring spikes outside the resected region relative to all detected spikes. We then reversed the training and validation centers and repeated the analysis. Predictive performance was quantified using AUROC and AUPRC. We hypothesized that the model developed using the larger cohort would yield better predictive performance.

### Standard protocol approvals, registrations, and patient consents

The institutional review boards of Wayne State University and the National Epilepsy Center, NHO Shizuoka Institute of Epilepsy and Neurological Disorders approved the study. Informed consent was obtained from guardians, and assent was obtained from participants aged 13 years or older.

## Results

### Patient characteristics

The cohort comprised 56 patients, including 36 treated in Shizuoka and 20 in Detroit (**Figure 2**; **Table 1**); 36 patients in Shizuoka and 11 in Detroit underwent a one-stage procedure because the clinical team judged that the resection target could be delineated using findings from the phase-1 noninvasive presurgical evaluation in conjunction with intraoperative ECoG. The remaining nine patients, all from Detroit, underwent a two-stage procedure including extraoperative iEEG recording. Mean age was 21.2 ± 13.2 years (range, 0.7–56.9 years), and 26 patients (46.4%) were female.

**Figure 2.**
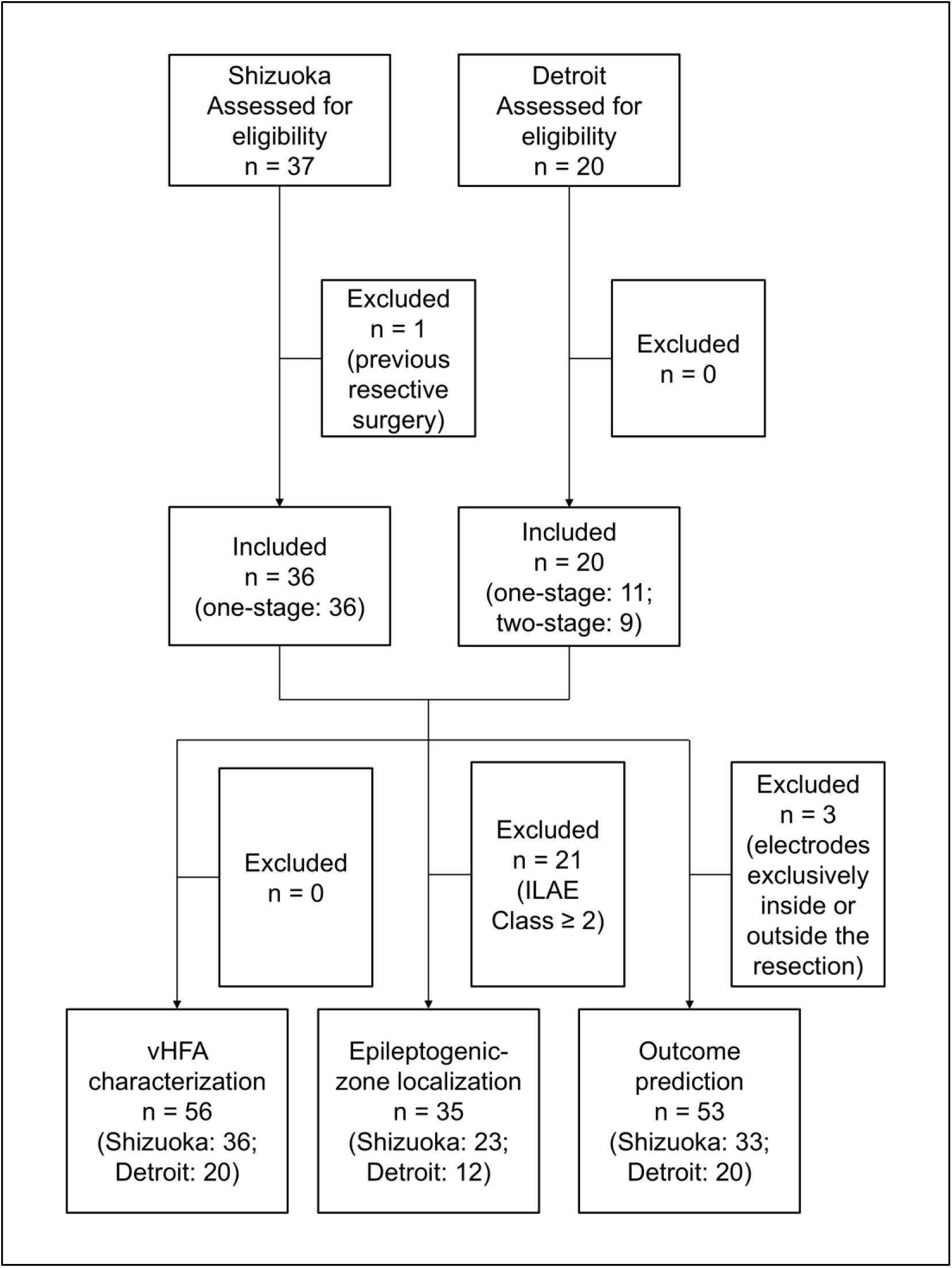
Patient Selection and Analytic cohorts. Abbreviations: ECoG = electrocorticography; ILAE = International League Against Epilepsy; vHFA = very high-frequency activity at 500–2,000 Hz

**Table 1.** Patient characteristics.

| Characteristic | Overall | Shizuoka | Detroit | Unadjusted p-value |
| --- | --- | --- | --- | --- |
| Number of patients | 56 | 36 | 20 |  |
| Age at surgery, years [range] | 21.2 ± 13.2 [0.7–56.9] | 27.0 ± 12.7 [6.4–56.9] | 10.8 ± 5.7 [0.7–17.6] | <0.001<sup>a</sup> |
| Female sex | 26/56 (46.4%) | 18/36 (50.0%) | 8/20 (40.0%) | 0.580<sup>b</sup> |
| Number of oral antiseizure medications | 2.1 ± 1.3 [0.0–6.0] | 2.4 ± 1.3 [1.0–6.0] | 1.6 ± 1.0 [0.0–3.0] | 0.041<sup>a</sup> |
| Daily seizures | 13/56 (23.2%) | 10/36 (27.8%) | 3/20 (15.0%) | 0.339<sup>b</sup> |
| Presence of MRI lesions | 53/56 (94.6%) | 35/36 (97.2%) | 18/20 (90.0%) | 0.288<sup>b</sup> |
| Number of intracranial electrodes | 38.4 ± 40.1 [8.0–134.0] | 14.8 ± 3.3 [8.0–18.0] | 80.7 ± 41.3 [16.0–134.0] | <0.001<sup>a</sup> |
| ECoG duration, min | 6.7 ± 2.9 [2.1–15.2] | 5.2 ± 1.1 [2.3–8.0] | 9.2 ± 3.4 [2.1–15.2] | <0.001<sup>a</sup> |
| Mean sevoflurane concentration, vol% | 1.9 ± 0.7 [1.0–3.1] | 1.5 ± 0.4 [1.0–3.1] | 2.6 ± 0.4 [1.8–3.0] | <0.001<sup>a</sup> |
| Two-stage surgery | 9/56 (16.1%) | 0/36 (0.0%) | 9/20 (45.0%) | <0.001<sup>b</sup> |
| Left-hemispheric surgery | 28/56 (50.0%) | 19/36 (52.8%) | 9/20 (45.0%) | 0.781<sup>b</sup> |
| Extratemporal resection | 30/56 (53.6%) | 18/36 (50.0%) | 12/20 (60.0%) | 0.580<sup>b</sup> |
| Follow-up duration, years | 5.3 ± 2.5 [1.0–9.8] | 5.3 ± 2.9 [1.0–9.8] | 5.3 ± 1.3 [1.9–7.0] | 1.000<sup>a</sup> |
| ILAE class 1 outcome | 35/56 (62.5%) | 23/36 (63.9%) | 12/20 (60.0%) | 0.781<sup>b</sup> |

Unadjusted P values are presented for comparisons of patient characteristics between Shizuoka and Detroit. <sup>a</sup> The Mann-Whitney U test was used for continuous variables. <sup>b</sup> The Fisher exact test was used for categorical variables. For sevoflurane concentration, patient-level mean values during ECoG were compared. ECoG = electrocorticography. ILAE = International League Against Epilepsy.

### Temporal features of vHFA

**eFigure 1** presents the distribution of artifact-free electrode contacts. The detector identified 132,359 spikes, of which 1,925 (1.45%) were vHFA-positive. Using patient-level median peak latencies, vHFA preceded the associated spike peak by 8.0 ms (95% CI, 5.0–16.0 ms; P<0.001).

### Factors associated with vHFA

Of 108,454 co-occurring spikes, 1,814 were vHFA-positive (399 of 30,813 leading spikes [1.29%] and 1,415 of 77,641 lagging spikes [1.82%]); in contrast, 111 of 23,905 isolated spikes (0.46%) were vHFA-positive. vHFA was more frequently associated with co-occurring spikes than with isolated spikes (OR: 3.65; 95% CI: 3.01–4.42; P<0.001). vHFA was less frequently associated with leading spikes than lagging spikes (OR: 0.71; 95%CI: 0.63–0.79; P<0.001).

Patients with daily seizures had higher odds of a given spike being vHFA positive than those without daily seizures (OR: 1.31; 95%CI: 1.18–1.46; P<0.001). Spikes from patients without MRI-visible lesions had higher odds of vHFA positivity than spikes from patients with MRI-visible lesions (OR: 1.43; 95%CI: 1.25–1.64; P<0.001). Spikes recorded from the medial temporal region had higher odds of being vHFA positive than those recorded outside this region (OR: 1.41; 95%CI: 1.24–1.61; P<0.001). A greater number of oral antiseizure medications (OR: 0.88; 95%CI: 0.85– 0.92; P<0.001) were associated with lower odds of a given spike being vHFA positive, whereas √age, sex, and sevoflurane concentration were not (P>0.17). Spikes recorded in Shizuoka had lower odds of being vHFA-positive than those recorded in Detroit (OR: 0.83; 95%CI: 0.73–0.93; P=0.002).

### Association between vHFA and spatial co-occurrence of spikes

vHFA-positive spike events had higher odds of being classified as co-occurring than vHFA-negative spike events, independent of medial temporal sampling and interelectrode distance (OR: 3.43; 95%CI: 2.82–4.17; P<0.001).

### Association between vHFA and the epileptogenic zone

Thirty-five of the 56 patients achieved ILAE class 1 outcomes. Among these 35 patients, intracranial electrode sites with higher rates of vHFA-positive co-occurring spikes were more likely to be located within the resected region (i.e., the presumed epileptogenic zone) (OR: 11.92 per occurrence/min; 95% CI: 5.62–25.32; P<0.001). This association was stronger than that observed for all spikes (OR: 1.09 per occurrence/min; 95%CI: 1.06–1.11; P<0.001). The strength of this association was comparable for that observed for HFA_250–500-Hz_ positive co-occurring spikes (OR: 9.20 per occurrence/min; 95%CI: 4.63–18.30; P<0.001). Associations of the remaining predictors with the resected region are provided in **Table 2**.

**Table 2.** Associations between spike types and the epileptogenic zone.

| **Spike type** | **OR per +1 occurrence/min**<br>**[95% CI]** | **Unadjusted**<br>**p-value** |
| --- | --- | --- |
| Spikes | 1.09 [1.06–1.11] | <0.001* |
| Co-occurring spikes | 1.12 [1.09–1.15] | <0.001* |
| vHFA-positive spikes | 10.17 [5.02–20.61] | <0.001* |
| vHFA-positive co-occurring spikes | 11.92 [5.62–25.32] | <0.001* |
| vHFA-positive leading spikes | 199.45 [22.58–1,761.42] | <0.001* |
| HFA<sub>250–500-Hz</sub> positive co-occurring spikes | 9.20 [4.63–18.30] | <0.001* |

Mixed-effects logistic regression evaluated whether contacts with higher rates of spike events were more likely localized within the resected brain region (i.e., the presumed epileptogenic zone) in 35 seizure-free patients. Each additional vHFA-positive co-occurring spike per minute was associated with approximately 11.92-fold higher odds of a contact being located within the presumed epileptogenic zone. \*: Significant after Bonferroni correction. OR = odds ratio. CI = confidence interval. HFA<sub>250–500-Hz</sub> = high-frequency activity at 250–500 Hz. vHFA = very high-frequency activity at 500–2,000 Hz.

### Association between vHFA and postoperative seizure recurrence

Logistic regression analysis of 53 patients revealed that a higher burden of vHFA-positive co-occurring spikes in preserved regions was associated with higher odds of postoperative seizure recurrence (OR: 3.22; 95% CI: 1.14–9.12; P=0.024; **Table 3**), although this association did not survive Bonferroni correction. This association was stronger than that observed for all detected spikes (OR: 1.00 per 0.01 increase in proportion; 95%CI: 0.98–1.02; P>0.05). Associations of the remaining predictors with seizure recurrence are provided in **Table 3**.

**Table 3.** Associations between spike types and postoperative seizure recurrence.

| Spike type | OR per 0.01 increase in proportion [95% CI] | Unadjusted p-value |
| --- | --- | --- |
| Spikes | 1.00 [0.98–1.02] | 0.993 |
| Co-occurring spikes | 1.00 [0.98–1.03] | 0.759 |
| vHFA-positive spikes | 2.95 [1.18–7.33] | 0.017 |
| vHFA-positive co-occurring spikes | 3.22 [1.14–9.12] | 0.024 |
| vHFA-positive leading spikes | 26.87 [1.36–530.98] | 0.027 |
| HFA<sub>250–500-Hz</sub> positive co-occurring spikes | 2.28 [0.97–5.34] | 0.051 |

Logistic regression evaluated whether a higher burden of each spike type in preserved regions was associated with postoperative seizure recurrence in 53 patients. None of the P values remained statistically significant after Bonferroni correction. CI = confidence interval. HFA<sub>250–500-Hz</sub> = high-frequency activity at 250–500 Hz. OR = odds ratio. vHFA = very high-frequency activity at 500–2,000 Hz.

### Prediction of postoperative seizure recurrence

In the Shizuoka-to-Detroit external validation, in which the prediction model based on vHFA-positive co-occurring spikes was developed using Shizuoka data and tested using Detroit data, the AUROC was 0.80 (95%CI: 0.56–1.00) and the AUPRC was 0.67 (95%CI: 0.36–0.83). In comparison, the model based on all detected spikes yielded an AUROC of 0.45 (95%CI: 0.20– 0.72) and an AUPRC of 0.35 (95%CI: 0.26–0.50). The DeLong test showed that the prediction model based on vHFA-positive co-occurring spikes had a higher AUROC than the model based on all detected spikes (P=0.005; **Figure 3**).

**Figure 3.**
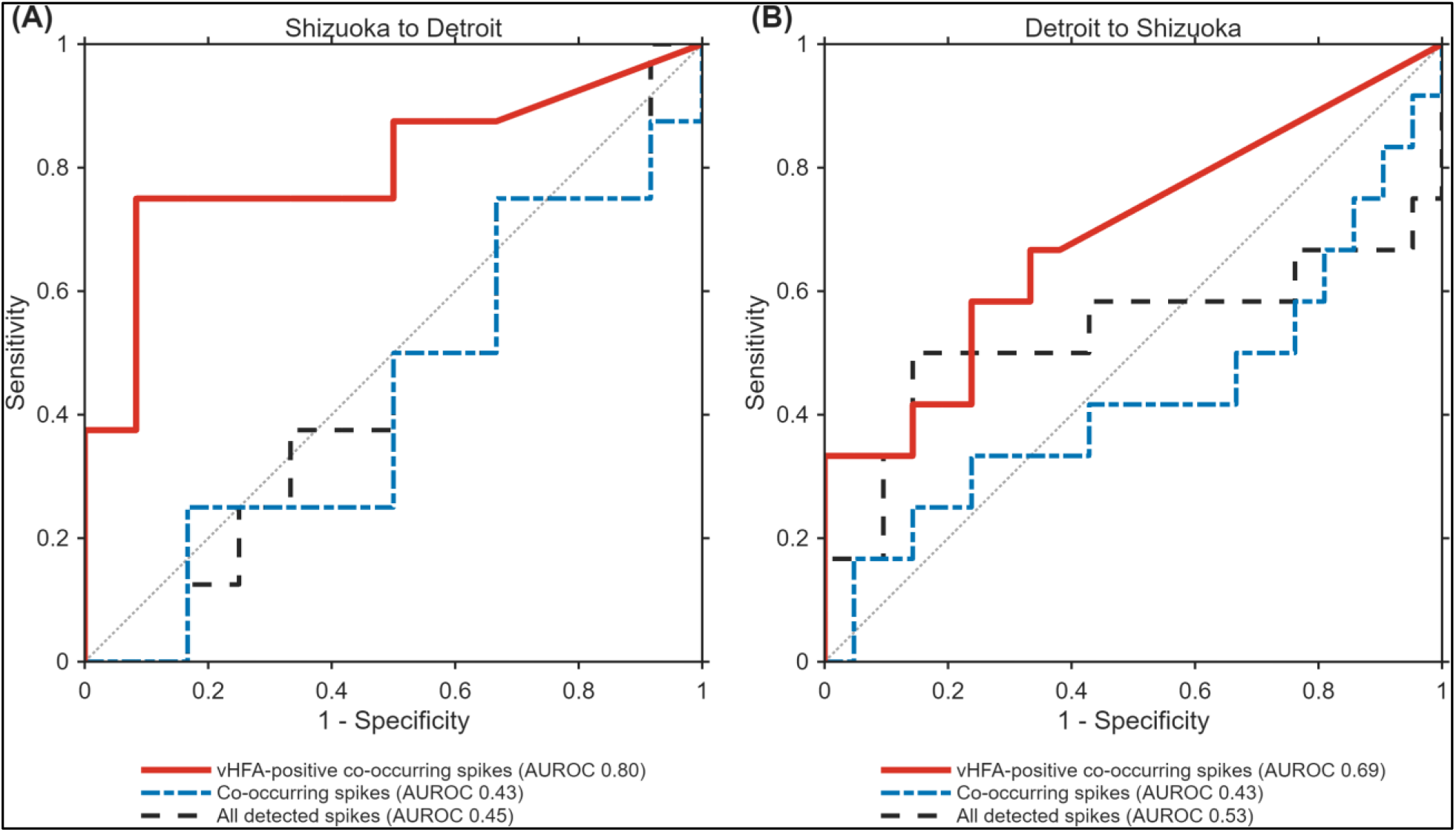
Area under the receiver operating characteristic curve (AUROC) for predicting postoperative seizure recurrence. **(A)** Cross-center external validation with models developed using Shizuoka data and tested in the independent Detroit cohort. **(B)** Cross-center external validation with models developed using Detroit data and tested in the independent Shizuoka cohort. vHFA = very high-frequency activity at 500–2,000 Hz.

In the Detroit-to-Shizuoka external validation, in which the prediction model based on vHFA-positive co-occurring spikes was developed using Detroit data and tested using Shizuoka data, the AUROC was 0.69 (95%CI: 0.50–0.87) and the AUPRC was 0.58 (95%CI: 0.32–0.73). In comparison, the model based on all detected spikes yielded an AUROC of 0.53 (95%CI: 0.28– 0.78) and an AUPRC of 0.46 (95%CI: 0.26–0.64). The prediction model based on vHFA-positive co-occurring spikes had a numerically higher AUROC than the model based on all detected spikes, although the difference was not statistically significant according to the DeLong test (P=0.462). Prediction performance for each spike type is provided in **Table 4**.

**Table 4.** Prediction of postoperative seizure recurrence.

| Spike type | AUROC<br>[95% CI] | AUPRC<br>[95% CI] |
| --- | --- | --- |
| **Shizuoka-to-Detroit external validation** |  |  |
| Spikes | 0.45 [0.20–0.72] | 0.35 [0.26–0.50] |
| Co-occurring spikes | 0.43 [0.18–0.70] | 0.35 [0.25–0.50] |
| vHFA-positive spikes | 0.77 [0.50–1.00] | 0.66 [0.34–0.82] |
| vHFA-positive co-occurring spikes | 0.80 [0.56–1.00] | 0.67 [0.36–0.83] |
| vHFA-positive leading spike | 0.70 [0.44–0.93] | 0.45 [0.30–0.69] |
| HFA<sub>250–500-Hz</sub> positive co-occurring spikes | 0.79 [0.53–0.99] | 0.68 [0.35–0.83] |
| **Detroit-to-Shizuoka external validation** |  |  |
| Spikes | 0.53 [0.28–0.78] | 0.46 [0.26–0.64] |
| Co-occurring spikes | 0.43 [0.21–0.65] | 0.35 [0.23–0.51] |
| vHFA-positive spikes | 0.68 [0.48–0.87] | 0.60 [0.33–0.75] |
| vHFA-positive co-occurring spikes | 0.69 [0.50–0.87] | 0.58 [0.32–0.73] |
| vHFA-positive leading spike | 0.72 [0.55–0.89] | 0.65 [0.38–0.78] |
| HFA<sub>250–500-Hz</sub> positive co-occurring spikes | 0.62 [0.43–0.81] | 0.46 [0.28–0.63] |

Cross-center validation assessed how well a model incorporating the abundance of each spike type in preserved regions, developed using data from one center, predicted postoperative seizure recurrence in the independent cohort from the other center. AUROC = area under the receiver operating characteristic curve. AUPRC = area under the precision-recall curve. CI = confidence interval. HFA<sub>250–500-Hz</sub> = high-frequency activity at 250–500 Hz. vHFA = very high-frequency activity at 500–2,000 Hz.

## Discussion

### Principal novel findings

To our knowledge, this is the first intraoperative ECoG study to characterize the spatiotemporal features of spike-associated vHFA and evaluate its prognostic value. The vHFA peak preceded the associated spike peak by a median of 8.0 ms, and spike-associated vHFA was approximately 3.7 times more likely to occur with spatially co-occurring spikes than with isolated spikes. Among patients who achieved ILAE class 1 outcomes, each one-per-minute increase in the rate of vHFA-positive co-occurring spikes was associated with approximately 12-fold higher odds of a contact being located within the presumed epileptogenic zone. This association was substantially stronger than that observed for all detected spikes. A higher burden of vHFA-positive co-occurring spikes in preserved regions was associated with postoperative seizure recurrence. Bidirectional cross-center validation supported the generalizability of this biomarker: a logistic regression model incorporating vHFA-positive co-occurring spikes developed using Shizuoka data predicted seizure recurrence in the independent Detroit cohort with an AUROC of 0.80, while the reverse validation also demonstrated predictive utility. These findings suggest that spike-associated vHFA identifies a biologically meaningful subset of interictal spikes involved in propagation across the epileptogenic zone and that vHFA-positive co-occurring spikes may serve as a generalizable intraoperative biomarker of the epileptogenic zone.

### Clinical significance of spike-associated vHFA

Our findings suggest that less than 10 minutes of intraoperative ECoG under sevoflurane may provide clinically useful information for localizing the epileptogenic zone. Sevoflurane has been reported to induce frequent, spatially distributed spikes resembling those observed during non-REM sleep,^21,31^ allowing identification of spike-associated vHFA and characterization of its spatial distribution. The strong enrichment of vHFA-positive co-occurring spikes within the resected region among patients who achieved seizure freedom suggests that this biomarker may help delineate the epileptogenic zone within the limited time available for intraoperative ECoG.

Intraoperative vHFA analysis of ECoG recorded under sevoflurane may be useful for centers performing one-stage resective surgery without extraoperative iEEG monitoring,^32,33^ where rapid delineation of the epileptogenic zone could help guide the extent of resection. Even when intraoperative ECoG is performed as part of a planned two-stage evaluation involving extraoperative iEEG, mapping the distribution of vHFA-positive co-occurring spikes could provide an opportunity to simulate resection of the presumed epileptogenic zone while the patient is still in the operating room. Such information could also help determine whether additional electrode sampling is warranted before completing the intraoperative evaluation. A recent study demonstrated the feasibility of real-time intraoperative HFA detection,^34^ and a similar framework will need to be developed for intraoperative detection and assessment of spike-associated vHFA. Prospective studies are needed to determine whether incorporating spike-associated vHFA into intraoperative decision-making improves surgical planning and postoperative seizure outcomes across geographically and methodologically diverse settings.

### Potential mechanisms underlying spike-associated vHFA

The biological mechanisms underlying spike-associated vHFA remain uncertain. The association between more complete resection of regions exhibiting vHFA-positive co-occurring spikes and postoperative seizure freedom suggests that spike-associated vHFA may represent pathologically excitatory neuronal activity within the epileptogenic zone. vHFA peaks preceded the associated spike peaks by a median of 8 ms, and vHFA was more frequently associated with spatially co-occurring than isolated spikes. These observations raise the possibility that the neuronal activity underlying vHFA may contribute to, rather than simply follow, spike generation and propagation across the epileptogenic zone.^35^ One potential mechanism is that bursts of neuronal action potentials occurring with submillisecond temporal dispersion within the epileptogenic zone^36,37^ generate broadband HFA detectable as increased vHFA power on ECoG. Previous studies have suggested that HFA may show increasing specificity for the epileptogenic zone with increasing spectral frequency, with HFA_250–500 Hz_ being more closely associated with epileptogenicity than HFA_80–250 Hz_ and vHFA_500–2,000 Hz_ potentially providing even greater specificity.^8,38,39^

### Methodological considerations

Spike-associated vHFA cannot be readily explained as filter-related ringing caused by the sharp transient of an interictal spike.^40^ If vHFA primarily reflected ringing generated by the spike waveform, its power peak would be expected to occur near the spike peak. In contrast, the vHFA power peak preceded the associated spike peak by a median of 8 ms, arguing against a simple explanation based on spike-induced filter ringing. Furthermore, our detection procedure was designed to exclude high-frequency activity that could be explained by extrapolation of the spectral power augmentation observed at 50–250 Hz into frequencies above 500 Hz. Together, these observations suggest that spike-associated vHFA may represent a biologically meaningful electrophysiological signal associated with a subset of interictal spikes rather than merely a filter-related artifact.

### Study limitations

Our findings do not establish that vHFA represents a distinct periodic oscillation within the 500– 2,000 Hz range. Our time-frequency analysis quantified power within consecutive 50-Hz frequency bins and therefore did not distinguish narrowband periodic activity from broadband or aperiodic spectral components.^41^ Thus, the observed increase in vHFA power could reflect transient broadband activity associated with population neuronal firing rather than oscillations at a specific frequency. Future analyses separating periodic and aperiodic spectral components, including characterization of the aperiodic spectral slope, may provide further insight into the neurophysiological mechanisms underlying spike-associated vHFA.

The present study was not designed to determine the optimal sevoflurane concentration for detecting spike-associated vHFA. Previous studies have shown that higher sevoflurane concentrations can increase interictal spike occurrence, particularly in epileptogenic zone, without necessarily altering the relative spatial distribution of epileptiform activity between epileptogenic and nonepileptogenic regions.^21,31^ An increase in spike occurrence is conceptually distinct from an increase in the probability that an individual spike is accompanied by vHFA. Our findings do not establish whether sevoflurane concentration affects the likelihood that a given spike is vHFA-positive. Further studies systematically varying sevoflurane concentrations in larger cohorts are warranted to determine their effects on both spike occurrence and spike-associated vHFA.

Our outcome analysis did not indicate that a model incorporating vHFA-positive leading spikes provided greater accuracy for predicting seizure recurrence than one incorporating all vHFA-positive co-occurring spikes. For example, when the model was developed using the Shizuoka cohort and externally tested in the Detroit cohort, the model incorporating vHFA-positive co-occurring spikes yielded an AUROC of 0.80, whereas that incorporating vHFA-positive leading spikes yielded an AUROC of 0.70. This finding may be partly attributable to the relative rarity of vHFA-positive leading spikes. Only approximately 20% of vHFA-positive co-occurring spikes were classified as leading, with the remainder classified as lagging. Consequently, event rates based exclusively on leading spikes may provide relatively sparse measurements, potentially limiting their ability to reliably characterize the spatial distribution of epileptogenic activity during short intraoperative ECoG recordings. Future studies may benefit from complementary approaches that provide continuous measures, such as the strength of phase-amplitude coupling between vHFA and delta waves,^17^ rather than relying exclusively on discrete event counts that may amount to only one or two events per minute at individual recording sites.

Another possibility is that the neuronal generator initiating a vHFA-positive co-occurring spike may not be located directly beneath the electrode showing the earliest spike peak. The true generator could instead be located between the leading and lagging recording sites, with differences in propagation time determining the observed temporal sequence across electrodes. Source-localization approaches that account for propagation through underlying white-matter pathways, such as dynamic tractography-based methods,^10^ may therefore help more accurately estimate the origin of propagating spike activity and potentially improve localization of the epileptogenic zone.

## Funding statement

This work was supported by the National Institutes of Health (NS064033 to E.A.), the Uehara Memorial Foundation Postdoctoral Fellowship (20210301 to H.U., and 202441017 to K.H.) and the Japan Society for the Promotion of Science (202560628 to H.U., and JP22J23281, JP22KJ0323, 202560576 to N.K.).

## Competing interests

The authors report no competing interests. We confirm that we have read the Journal’s position on issues involved in ethical publication and affirm that this report is consistent with those guidelines.

## Supplementary material

Supplementary material includes **eFigure 1**.

## Author contributions

Study design: H.U., N.K., N.U., E.A.; Data acquisition: H.U., N.K., N.U., H.O., T.O., M.H., N.K., K.H., E.E., K.F., D.C., N.M., M.F., N.K., A.F.L., M.C., E.A.; Data analysis and interpretation: H.U., N.K., N.U., E.A.; Writing original draft: H.U., E.A.; Writing review and editing: E.A.; Supervision: E.A.

## Data Availability

All data produced in the present study are available upon reasonable request to the authors.

## Supplementary material

**eFigure 1a.**
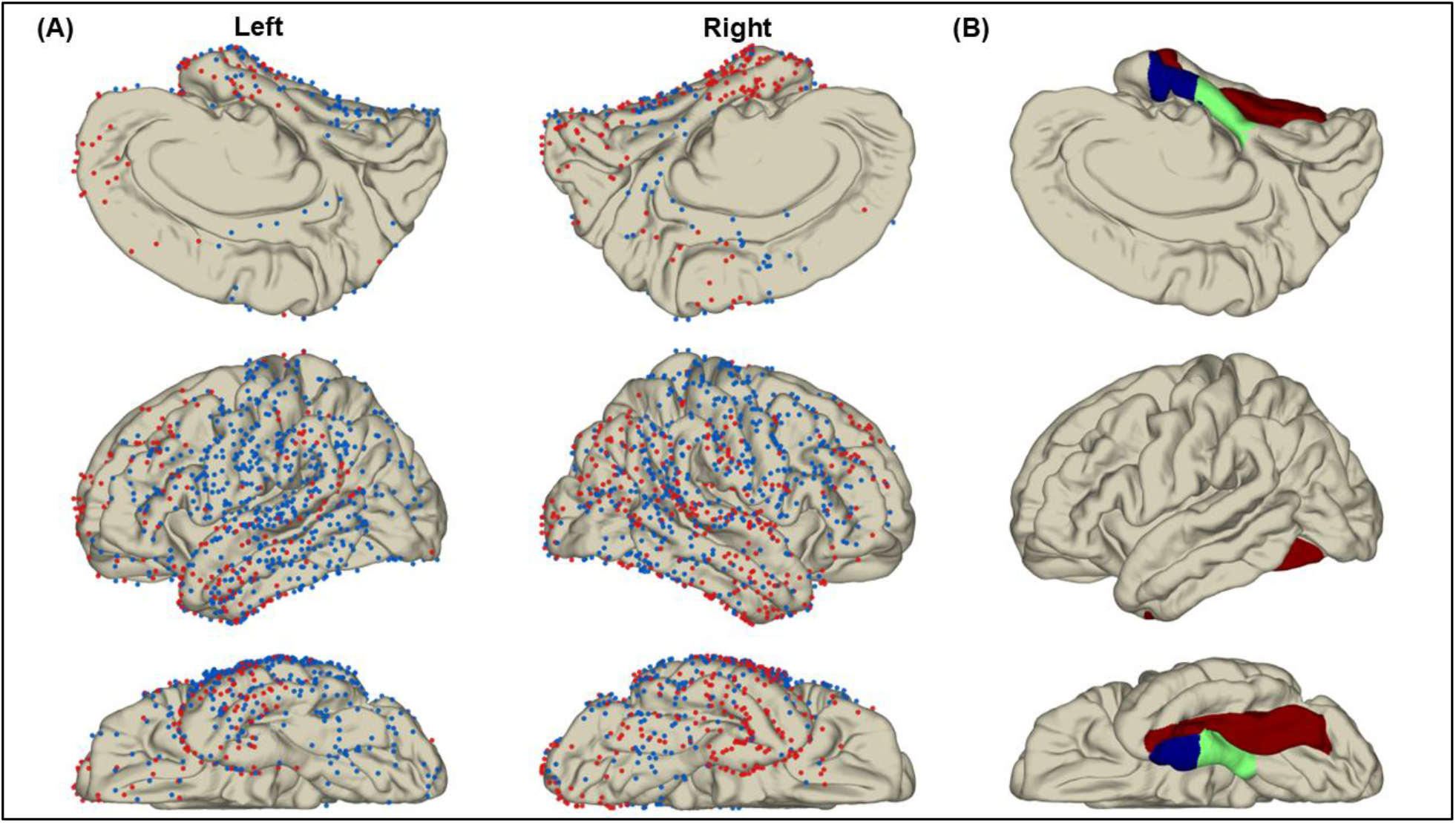
Distribution of artifact-free electrode contacts and medial temporal regions. **(A)** Artifact-free electrode contacts from 56 patients were projected onto the FreeSurfer average cortical surface (fsaverage). Blue and red dots indicate contacts outside and within the resected regions, respectively. The left and right hemispheres are shown in medial (top), lateral (middle), and inferior (bottom) views. **(B)** Anatomical regions used to define medial temporal sites. Blue, green, and brown indicate the entorhinal, parahippocampal, and fusiform regions, respectively.

## Notes

### Competing Interest Statement

The authors have declared no competing interest.

### Author Declarations

The Institutional Review Board of Wayne State University and the Ethics Committee of the National Epilepsy Center, NHO Shizuoka Institute of Epilepsy and Neurological Disorders independently approved this retrospective observational study.

